# Metabolic Hormone and Adipokine Alterations in Major Depressive Disorder in Relation to the Acute-Phase Inflammatory Response and Early-Life Adversity

**DOI:** 10.64898/2026.01.28.26345089

**Authors:** Tangcong Chen, Yueyang Luo, Mengqi Niu, Mengdie Li, Abbas F. Almulla, Marta Kubera, Yingqian Zhang, Michael Maes

## Abstract

Major depressive disorder (MDD) involves dysregulated neuroimmune, metabolic, and oxidative stress (NIMETOX) pathways. Recently, it was shown that NIMETOX pathways should be evaluated in MDD patients stratified for metabolic syndrome (MetS). The current study aims to characterize the metabolic hormone and adipokine profiles of Chinese MDD patients stratified for MetS and to delineate their associations with overall severity of depression (OSOD), suicidal ideation (SI), recurrence of illness (ROI), and physiosomatic symptoms. We enrolled 125 MDD inpatients and 40 healthy controls and measured fasting serum insulin, glucose, glucagon, Glucose-dependent Insulinotropic Polypeptide (GIP), Glucagon-Like Peptide-1 (GLP-1), leptin, secretin, Plasminogen Activator Inhibitor-1 (PAI-1), resistin, ghrelin, and adiponectin, as well as the acute-phase inflammatory (API) response using albumin, transferrin (Tf), and monomeric CRP (mCRP). The results revealed a distinct metabolic hormone and adipokine signature in MDD with significantly lower insulin, glucagon, and PAI-1 levels, alongside an elevated API index (after adjusting for age, MetS, and body mass index). A composite GAP index (ghrelin, adiponectin, PAI-1) correlated negatively with OSOD, SI, ROI, physiosomatic symptoms, and adverse childhood experiences (ACEs). Integrative modeling combining the GAP index, API index, and ACEs achieved an area under the receiver operating characteristic (ROC) curve of 0.864 with an accuracy of 80% for discriminating MDD from controls. In conclusion, the findings delineated that many inpatients with severe MDD suffer from suppressed anabolic hormones and lower adipokine levels coupled with a mild, chronic inflammatory response. The deviations in this “hormonal-immune-metabolic” axis are components of the NIMETOX pathways in MDD and are not associated with MetS.

## Introduction

Major depressive disorder (MDD) is increasingly reconceptualized as a systemic disorder involving dysregulation of neuroimmune, metabolic, and oxidative stress (NIMETOX) pathways ^1^. A major neuro-immune marker in MDD is the presence of an acute-phase inflammatory (API) response as indicated by lower serum albumin and transferrin (two negative acute-phase proteins) and increased monomeric C-reactive protein (mCRP), a positive acute phase response^2^. Combining these three markers into a single composite score (the API index) increases accuracy for MDD^2^. Increased levels of pro-inflammatory cytokines in MDD, such as interleukin (IL)-1, IL-6, and tumor necrosis factor-alpha (TNF-α), induce the API response^3^.

Growing evidence indicates that MDD is consistently accompanied by a pro-atherogenic lipid signature—lower high-density lipoprotein cholesterol (HDL), apolipoprotein (Apo)A1, and a diminished index of reverse cholesterol transport (RCT) —observed across multiple populations^4,5^. Our results show that in Chinese patients without MetS/obesity, these deficits persist after adjusting for the API response, confirming lipid dysregulation is intrinsic to MDD ^6^.

Within the NIMETOX framework, metabolic disorders, including metabolic syndrome (MetS) represent a pathological link between peripheral NIMETOX pathways and brain dysfunction (Nunes et al., 2015; Maes, Almulla, You, et al.,^1,7^. MetS is characterized by insulin resistance (IR) and increased atherogenicity, comprising a cluster of metabolic abnormalities including central obesity, dyslipidemia, hypertension, and impaired glucose metabolism that collectively promote cardiovascular disease risk ^8^. Conversely, depressive states may exacerbate metabolic dysfunction through neuroendocrine and behavioral pathways, suggesting a bidirectional relationship ^9^.

However, empirical evidence regarding IR in MDD is more heterogeneous than increased atherogenicity. A meta-analysis by Fernandes et al. demonstrates that IR is increased in MDD, particularly in Western studies^10^. However, these findings lack adjustment for MetS status, highlighting the need for more nuanced investigations that disentangle primary metabolic disturbances in MDD from those secondary to comorbid MetS. Thus, after adjusting for MetS and body mass index (BMI), such differences were not always evident ^8,11^. While large-scale studies, including in Chinese cohorts, show that elevated IR markers like the triglyceride-glucose (TyG) index predict depression risk^12^, a study by Luo et al. revealed that Chinese patients with MDD—after controlling for MetS—had lower fasting insulin and IR index than controls^13^. This report identified MetS as a critical effect modifier: in its absence, mild inflammation in MDD may induce a context-dependent enhancement of insulin sensitivity; in its presence, pre-primed inflammation exacerbates IR ^13,14^. This underscores the need for metabolically stratified analyses rather than blanket assertions of a positive MDD-IR correlation.

The NIMETOX disturbance in MDD extends beyond classic hormones to a broader “immune-metabolic-endocrine axis,” involving metabolic hormones and adipokines that communicate between metabolic tissues, the immune system, and the brain ^1,15^. These hormonal mediators are pivotal in linking metabolic dysregulation to core clinical outcomes. For instance, ghrelin, an orexigenic hormone, has been shown to possess antidepressant and anti-inflammatory properties ^16,17^. Its altered levels in MDD are linked to changes in appetite and modulation of the stress response. Plasminogen Activator Inhibitor-1 (PAI-1), a pro-inflammatory adipokine and marker of IR, is paradoxically lower in some MDD patients, associated with a more severe inflammatory state ^18^. Adiponectin, an insulin-sensitizing, anti-inflammatory adipokine, is often reduced in MDD, correlating with increased severity and inflammation ^19,20^. Other hormones like glucagon, GLP-1, and leptin further contribute to this dysregulated network, affecting energy homeostasis and neuroplasticity ^21,22^. Their collective dysfunction suggests a distinct metabolic-endocrine subtype of depression ^23^.

Clinically, MDD is multifaceted. Bifactor modeling reveals that the symptoms load onto a general factor (overall severity of depression, OSOD) and a single group physiosomatic factor^24,25^. A key feature of the illness trajectory of MDD is the recurrence of illness (ROI), i.e., the recurrence of lifetime episodes and suicidal behaviors^26,27^. ROI is strongly predicted by ACEs and mediates the effects of ACEs on worsening OSOD, SI, and physiosomatic symptoms^28,29^. This ACE-ROI axis is linked to progressive sensitization of NIMETOX pathways, including inflammation and oxidative stress ^30–32^.

Despite this, the specific interrelationships between hormonal-metabolic markers (e.g., ghrelin, PAI-1, adiponectin) and these key clinical constructs remain poorly delineated, especially in Chinese populations. Current knowledge relies heavily on Western data, overlooking ethnic variations and differences in body composition (e.g., visceral adiposity), and lifestyle ^33^. Consequently, research mapping the relationships between IR, metabolic hormones, inflammation, and multidimensional clinical features in Chinese MDD is urgently needed.

Hence, we conduct a study to investigate the alterations in metabolic hormones and adipokines among Chinese MDD patients, and to examine their associations with the core clinical dimensions of MDD, namely OSOD, current SI, ROI, and physiosomatic symptoms. In addition, we explore the associations between hormonal/metabolic markers and the API response in MDD.

## 2. Methods

### 2.1. Study Design and Participants

This cross-sectional case-control study enrolled 165 participants, including 125 inpatients meeting DSM-5 criteria for MDD and 40 age-, sex-, BMI-, and education-matched healthy controls. Participants were recruited from the Psychiatric Center of Sichuan Provincial People’s Hospital in Chengdu, China. MDD diagnosis was confirmed using the Mini-International Neuropsychiatric Interview (MINI), with a severity threshold of >18 on the 21-item Hamilton Depression Rating Scale (HAMD-21).

Exclusion criteria for both groups comprised medical disorders, such as diabetes type 1, immune and autoimmune disorders, including inflammatory bowel disease, rheumatoid arthritis, neurological and neurodegenerative disorders, such as Parkinson’s and Alzheimer’s disease, multiple sclerosis), or psychiatric disorders, such as bipolar disorder, schizophrenia, substance use disorders, autism spectrum disorders), infectious disease, immunomodulatory treatments, treatments with therapeutical dosages of antioxidants or omega-3 polyunsaturated fatty acids (Niu et al., 2025). We also excluded pregnant and lactating women. Controls were additionally excluded for MDD and a family history of mood disorders, suicide or psychosis.

The study was approved by the ethics committee of Sichuan Provincial People’s Hospital ([Ethics (Research) 2024-203]), and all participants provided written informed consent.

### 2.2. Clinical and Behavioral Assessments

Trained clinicians conducted semi-structured interviews to collect demographic and clinical data. **Supplementary Table 1** shows the instruments we employed to assess the clinical indices as described before^25^. Based on those measurements, composite clinical constructs were generated as previously described to construct OSOD, current suicidal ideation (Current SI), ROI, and physiosomatic symptoms. ACEs were quantified using the Childhood Trauma Questionnaire-Short Form^34^.

### 2.3. Anthropometric and Metabolic Syndrome Assessment

BMI was calculated using the measured height and weight. Waist circumference (WC) was measured at the midpoint between the iliac crest and the lowest rib. MetS was defined according to the 2009 Joint Interim Statement criteria^35^, requiring at least three of five components: elevated WC, triglycerides, blood pressure, fasting glucose, or reduced HDL-cholesterol.

### 2.4. Assays

Fasting venous blood was collected in the morning using EDTA-containing tubes. Following centrifugation, plasma was aliquoted and stored at –80°C until analysis. Fasting plasma glucose (FPG) was measured using the glucose oxidase method on a fully automated biochemistry analyzer (ADVIA 2400, Siemens Healthcare Diagnostics Inc.). Insulin was assayed using a chemiluminescence immunoassay (Atellica IM insulin assay kit, Siemens Healthcare Diagnostics Inc.) on an Atellica IM analyzer. To assess insulin resistance, we computed a composite indicator (zFPG + zINS) by summing the standardized z-scores of fasting plasma glucose and insulin levels ^36^. The API response was assessed by measuring plasma levels of albumin, transferrin (Tf), and monomeric C-reactive protein (mCRP). The mCRP values were adjusted for the effects of age, sex, and BMI, and the residualized mCRP values were used in subsequent analyses^37^. The API index was then calculated using the formula: z mCRP – z albumin – z transferrin.

Metabolic hormones (Ghrelin, GIP, GLP-1, Glucagon, Leptin, Secretin) and adipokines (Resistin, PAI-1, Adiponectin) were quantified using multiplex magnetic bead-based immunoassays (MILLIPLEX® Human Metabolic Hormone Panel V3 and Human Adipokine Magnetic Bead Panel 1, respectively) based on Luminex® xMAP® technology. Assays were performed according to the manufacturer’s protocols, including preparation of standards and controls, plate washing, and signal acquisition on a Luminex instrument. The intra-assay coefficients of variation for all analytes were <10%, respectively. Data were analyzed using a 5-parameter logistic curve-fitting method to determine analyte concentrations.

### 2.5. Statistical Analysis

Statistical analyses were performed using SPSS 30.0 (IBM Corp., Armonk, NY, USA). Continuous data are presented as mean ± standard deviation, and categorical data as frequencies (percentages). Group differences (MDD vs. controls) in continuous variables were tested using Analysis of Variance (ANOVA) or the General Linear Model (GLM). GLM was employed to compare hormonal and adipokine biomarkers between groups while adjusting for the effects of age, sex, BMI, and MetS status, with results presented as estimated marginal means. Associations between categorical variables were assessed using the chi-square test. For all group comparisons, the False Discovery Rate (FDR) procedure was applied to adjust p-values for multiple testing.

Associations between continuous variables were examined using Pearson’s correlation analysis. To identify predictors of clinical outcomes (OSOD, ROI, Current SI, physiosomatic symptoms), multiple linear regression analyses were conducted. Independent variables, including ACEs, hormonal biomarkers, and inflammatory indices, were entered into the model using stepwise selection. Model assumptions, including linearity, homoscedasticity, independence of residuals, and absence of multicollinearity, were checked and met. Binary logistic regression analysis was performed to construct diagnostic models differentiating MDD patients from healthy controls. Significant variables were then entered into the multivariate models using the forward conditional method. The model’s discriminatory power was evaluated by the area under the curve (AUC) of the receiver operating characteristic (ROC) curve. Model fit was assessed with the Hosmer-Lemeshow test, and the overall classification accuracy, sensitivity, and specificity were reported. Odds ratios (OR) with 95% confidence intervals (CI) were calculated for each significant predictor. The diagnostic performance of key biomarker combinations was further evaluated using ROC analysis. A two-tailed alpha level of 0.05 was considered statistically significant for all tests unless otherwise specified.

## 3. Results

### 3.1 Sociodemographic and Basic Clinical Characteristics

**Table 1** presents the sociodemographic and basic clinical profiles of patients and controls. No statistically significant differences were observed between the two groups in age, sex, BMI, WC, years of education, prevalence of MetS, or smoking history. Patients with MDD showed higher OSOD, ROI, SI, physiosomatic, and ACE scores than controls.

**Table 1.** Clinical, socio-demographic, and biochemical data of patients with major depressive disorder (MDD) and healthy controls (HC)

| Variables | HC (n=40) | MDD (n=125) | F/$\chi^2$ | df | p |
| --- | --- | --- | --- | --- | --- |
| Age (years) | 37.1(13.8) | 35.7 (12.1) | 0.37 | 1/163 | 0.542 |
| Gender (m/f) | 13/27 | 38/87 | 0.06 | 1 | 0.802 |
| BMI (kg/m<sup>2</sup>) | 23.52 (4.07) | 22.30 (3.6) | 3.58 | 1/163 | 0.06 |
| Waist circumference (cm) | 79.38 (11.73) | 78.27 (11.32) | 0.28 | 1/163 | 0.595 |
| Ranking MetS | 1.590 (1.46) | 1.42 (1.26) | 0.5 | 1/161 | 0.479 |
| Education years (years) | 13.88 (4.34) | 13.54 (3.33) | 0.26 | 1/163 | 0.613 |
| Metabolic syndrome (MetS) (No/Yes) | 29/10 | 102/22 | 1.17 | 1 | 0.355 |
| History of smoking (No/Yes) | 37/3 | 101/24 | 3.03 | 1 | 0.091 |
| OSOD | -1.518 (0.237) | 0.528 (0.491) | 640.21 | 1/153 | <0.001 |
| Four ACEs | -0.785 (0.170) | 0.150 (0.110) | 22.87 | 1/155 | <0.001 |
| Physiosomatic | -1.316 (0.120) | 0.513 (0.091) | 158.22 | 1/139 | <0.001 |
| Current SI | -0.687 (0.163) | 0.222 (0.117) | 15.419 | 1/145 | <0.001 |
| ROI | -1.032 (0.140) | 0.274 (0.101) | 31.793 | 1/145 | <0.001 |

All results are shown as mean (SD).
BMI: Body Mass Index, OSOD: Overall Severity of Depression, ACEs: Adverse Childhood Experiences, Current SI: Current Suicidal Ideation, ROI: Reoccurrence of illness index.

### 3.2 Distinct Neuroendocrine and Metabolic Alterations in MDD

**Table 2** shows the significant alterations in hormonal and metabolic profiles observed in MDD patients after adjusting for age and BMI (sex was not significant). Key findings include a significant reduction in fasting insulin, glucagon, and PAI-1 levels. Concurrently, the API index was markedly elevated in the MDD group. No significant intergroup differences were found for GIP, GLP-1, leptin, secretin, resistin, or adiponectin.

**Table 2.** Differences in hormone profiles between major depressive disorder (MDD) and healthy controls (HC)

| Variables | HC (n=40) | MDD (n=125) | F/$\chi^2$ | df | P | Partial Eta Squared |
| --- | --- | --- | --- | --- | --- | --- |
| FPG (mmol/L) | 5.563 (0.086) | 5.374 (0.056) | 3.841 | 1/155 | 0.109 | 0.023 |
| Insulin (mU/L) | 8.792 (0.586) | 6.559 (0.380) | 10.967 | 1/155 | 0.005 | 0.066 |
| API index | -1.437 (0.346) | 0.498 (0.225) | 23.560 | 1/155 | <0.001 | 0.132 |
| Ghrelin (unmeasurable/measurable) | 32/7 | 119/6 | 7.041 | 1 | 0.038 | - |
| GIP (pg/ml) | 70.193 (6.678) | 61.319 (4.336) | 1.333 | 1/155 | 0.288 | 0.009 |
| GLP-1 (pg/ml) | 32.769 (7.417) | 17.153 (4.816) | 3.345 | 1/155 | 0.110 | 0.021 |
| Glucagon (pg/ml) | 49.168 (3.761) | 37.546 (2.468) | 9.556 | 1/155 | 0.008 | 0.058 |
| Leptin (ng/ml) | 5.288 (0.858) | 5.111 (0.557) | 0.032 | 1/155 | 0.858 | <0.001 |
| Secretin (pg/ml) | 56.078 (8.500) | 36.212 (5.519) | 4.122 | 1/155 | 0.094 | 0.026 |
| Resistin (ng/ml) | 2.615 (0.347) | 1.895 (0.225) | 3.252 | 1/155 | 0.110 | 0.021 |
| PAI-1 (ng/ml) | 3.329 (0.465) | 1.795 (0.305) | 8.163 | 1/131 | 0.015 | 0.059 |
| Adiponectin (ng/ml) | 10.718 (1.479) | 9.315 (0.961) | 0.679 | 1/155 | 0.440 | 0.004 |

All results are shown as estimated marginal means (SE), after adjusting for age and BMI.
FPG: Fasting Plasma Glucose, API: Acute Phase Inflammatory, GLP-1: Glucagon-Like Peptide-1, PAI-1: Plasminogen Activator Inhibitor-1.

### 3.3 Results of Binary Logistic Regression

To assess the accuracy of the aforementioned biomarkers, a series of binary logistic regression models was constructed (**Table 3**, **Supplementary Table 2**). The discriminatory performance of the models improved progressively as variables from different pathophysiological domains were integrated: a baseline model containing only hormonal markers (ghrelin, PAI-1, adiponectin) yielded an AUC of 0.741. Based on this model, we constructed a z-unit-based adipokine composite score as z PAI-1 + z adiponectin + ghrelin (0 vs 1), labeled the GAP index (the first letter of the markers). The inclusion of a metabolic marker (FPG) did not significantly improve performance; the addition of the inflammatory marker (API index) increased the AUC to 0.776, and further incorporation of sex and ACEs resulted in an AUC of 0.818. The optimal diagnostic model was constructed by integrating the GAP index, API index, and ACEs (Model 5). This model demonstrated adequate discriminatory power: AUC = 0.864 (SE = 0.031), with an overall accuracy of 80.0% (sensitivity 79.8%, specificity 74.4%). The corresponding ROC curve is shown in **Figure 1**.

**Figure 1.**
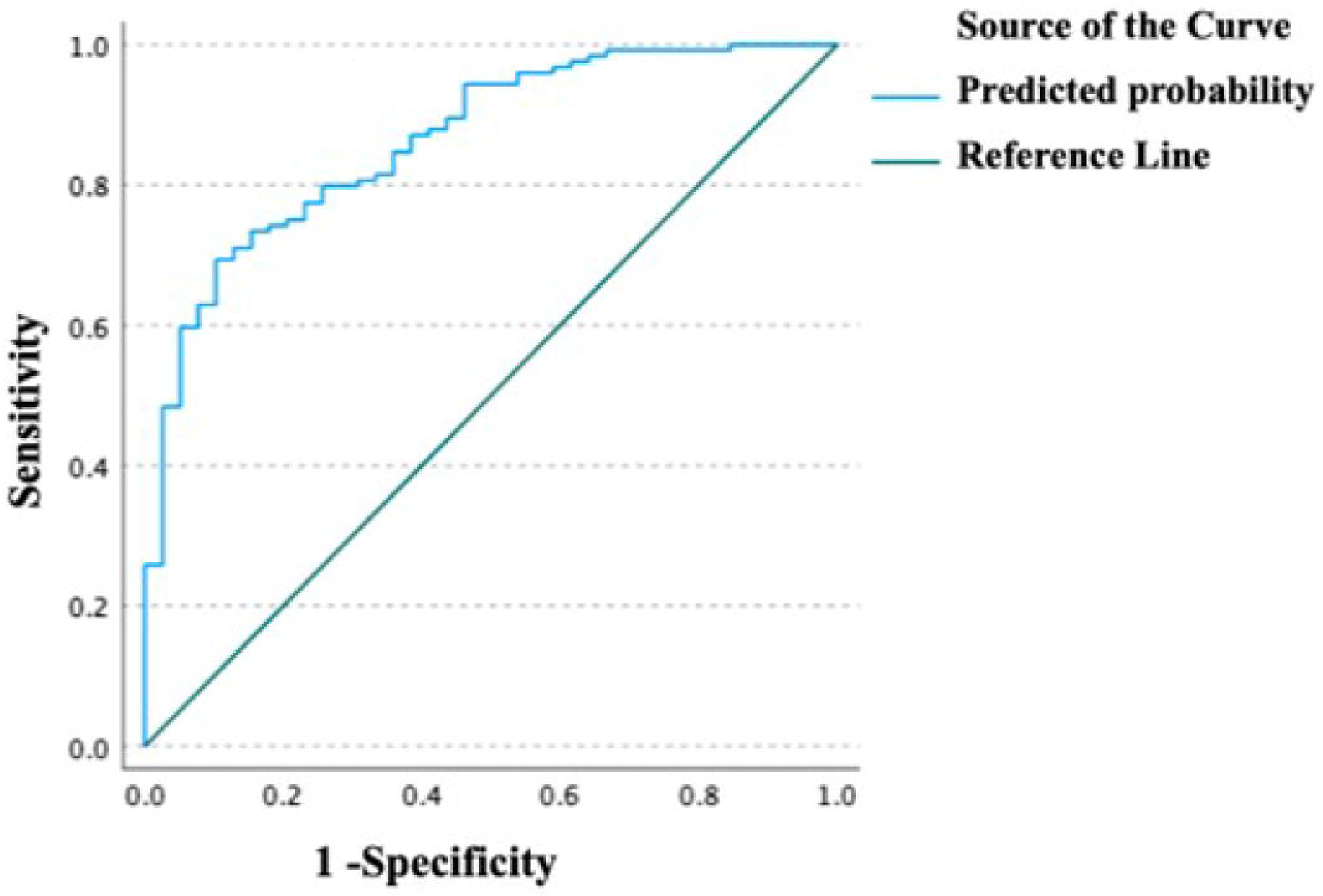
The receiving operating curve discriminating major depressive disorder from controls using GAP index, API index, and Four ACEs as explanatory variables.

**Table 3.**
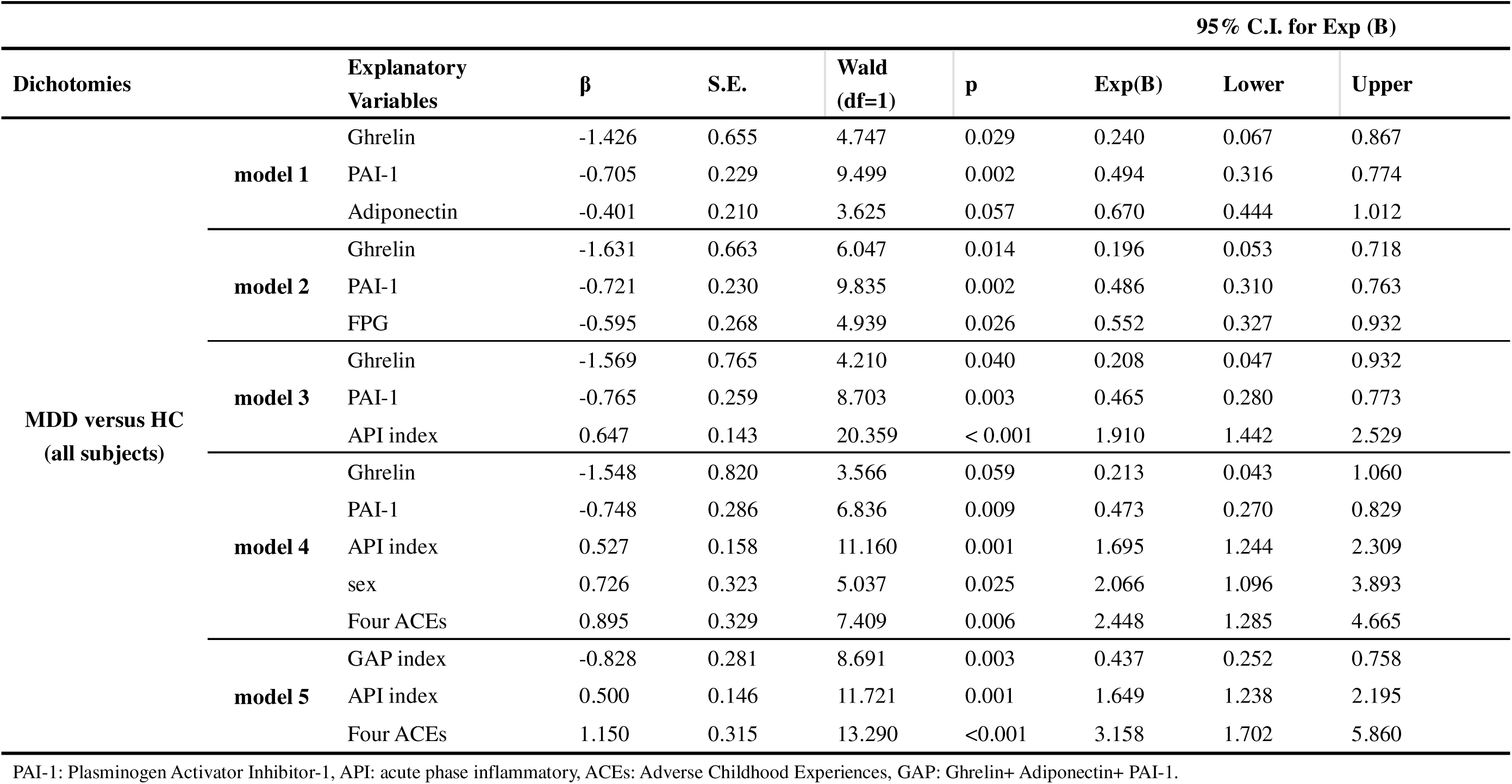
Results of binary logistic regression analysis examining the discrimination of major depressive disorder (MDD) and healthy controls (HC)

### 3.4 Correlations among Hormonal, Metabolic, and Inflammatory Markers

Pearson correlation analysis in **Table 4** shows the relationships between key variables. The API index showed significant negative correlations with PAI-1, adiponectin, and the GAP composite index. Concurrently, PAI-1, adiponectin, and the GAP index were significantly positively correlated with FPG, insulin, and the IR index (z FPG + z INS).

**Table 4.** Intercorrelation matrix (Pearson’s correlation coefficients) between hormone profile, insulin biomarkers, and metabolic indicators.

| Variables | Four_ACEs | API index | FPG | Insulin | z FPG + z INS |
| --- | --- | --- | --- | --- | --- |
| **Ghrelin** | -0.053 | -0.118 | 0.042 | 0.012 | 0.032 |
| **GIP** | -0.099 | 0.044 | 0.144 | 0.182* | 0.189* |
| **GLP-1** | -0.111 | -0.027 | 0.114 | 0.024 | 0.080 |
| **Glucagon** | 0.038 | -0.089 | 0.141 | 0.147 | 0.167* |
| **Leptin** | 0.025 | -0.023 | 0.136 | 0.277** | 0.240** |
| **Secretin** | -0.018 | 0.033 | 0.168* | -0.064 | 0.061 |
| **PAI-1** | -0.204** | -0.189* | 0.298** | 0.217** | 0.299** |
| **Adiponectin** | -0.217** | -0.319** | 0.126 | 0.216** | 0.198* |
| **Resistin** | -0.037 | -0.170* | 0.131 | -0.022 | 0.063 |
| **GAP index** | -0.270** | -0.340** | 0.274** | 0.274** | 0.318** |

\*\* Correlation is significant at the 0.01 level; \* Correlation is significant at the 0.05 level.
ACEs: Adverse Childhood Experiences, API: Acute Phase Inflammatory, FPG: Fasting Plasma Glucose, INS: Insulin, GIP: Glucose-dependent Insulinotropic Polypeptide, GLP-1: Glucagon-Like Peptide-1, PAI-1: Plasminogen Activator Inhibitor-1, GAP: zGhrelin+ zAdiponectin+ zPAI-1.

### 3.5 Hormone Profiles, Adipokines, and Clinical Symptom Severity

**Table 5** presents correlations between hormonal/metabolic indicators and clinical symptoms. Ghrelin, PAI-1, adiponectin, FPG, and the IR index were all significantly negatively correlated with OSOD, physiosomatic symptoms, and ROI. Among these, the GAP composite index showed the strongest negative correlations with all clinical dimensions.

**Table 5.** Intercorrelation matrix (Pearson’s correlation coefficients) between hormone profile and severity scales.

| Variables | Current SI | OSOD | Physiosomatic symptoms | ROI | Weight loss |
| --- | --- | --- | --- | --- | --- |
| **Ghrelin** | -0.053 | -0.211<sup>*</sup> | -0.13 | -0.210<sup>**</sup> | -0.212<sup>**</sup> |
| **GIP** | -0.099 | -0.114 | -0.104 | -0.139 | -0.041 |
| **GLP-1** | -0.111 | -0.083 | -0.008 | -0.127 | -0.097 |
| **Glucagon** | 0.038 | -0.026 | -0.003 | -0.059 | -0.019 |
| **Leptin** | 0.025 | -0.097 | -0.058 | -0.049 | -0.193<sup>*</sup> |
| **Secretin** | -0.018 | -0.070 | -0.001 | -0.129 | -0.081 |
| **PAI-1** | -0.204<sup>**</sup> | -0.316<sup>**</sup> | -0.246<sup>**</sup> | -0.199<sup>*</sup> | -0.185<sup>*</sup> |
| **Adiponectin** | -0.217<sup>**</sup> | -0.220<sup>**</sup> | -0.220<sup>**</sup> | -0.159<sup>*</sup> | -0.128 |
| **Resistin** | -0.037 | -0.194<sup>*</sup> | -0.172<sup>*</sup> | -0.234<sup>**</sup> | -0.093 |
| **Insulin** | -0.110 | -0.200<sup>*</sup> | -0.187<sup>*</sup> | -0.201<sup>*</sup> | -0.118 |
| **FPG** | -0.216<sup>**</sup> | -0.227<sup>**</sup> | -0.185<sup>*</sup> | -0.277<sup>**</sup> | -0.186<sup>*</sup> |
| **z FPG + z INS** | -0.189<sup>*</sup> | -0.246<sup>**</sup> | -0.214<sup>**</sup> | -0.277<sup>**</sup> | -0.176<sup>*</sup> |
| **GAP index** | -0.270<sup>**</sup> | -0.378<sup>**</sup> | -0.318<sup>**</sup> | -0.266<sup>**</sup> | -0.238<sup>**</sup> |

\*\* Correlation is significant at the 0.01 level; \* Correlation is significant at the 0.05 level.
Current SI: Current Suicidal Ideation, OSOD: Overall Severity of Depression, API: Acute Phase Inflammatory, FPG: Fasting Plasma Glucose, INS: Insulin, GIP: Glucose-dependent Insulinotropic Polypeptide, GLP-1: Glucagon-Like Peptide-1, PAI-1: Plasminogen Activator Inhibitor-1, GAP: Ghrelin+ Adiponectin+ PAI-1, ROI: Reoccurrence of illness index.

Further multiple regression analysis (**Table 6**) quantified the contribution of these predictors. ACEs and the API index were the most consistent and significant positive predictors for OSOD, physiosomatic symptoms, and illness recurrence. Additionally, specific hormonal markers had independent predictive value: for example, ghrelin and PAI-1 were negative predictors of OSOD, while resistin and GLP-1 were negative predictors of current SI. These models collectively explained between 30.1% and 40.2% of the variance in clinical symptom severity.

**Table 6.**
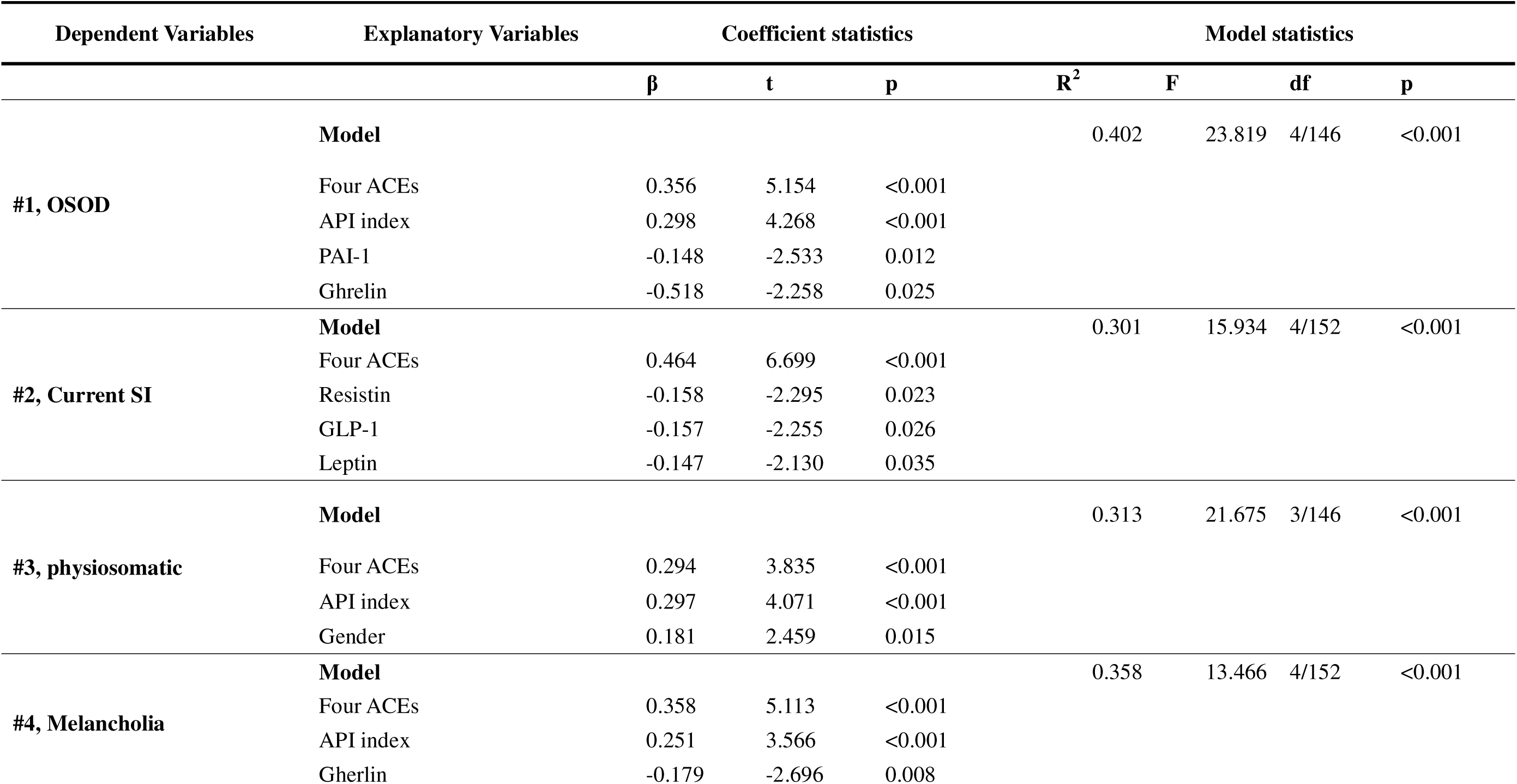
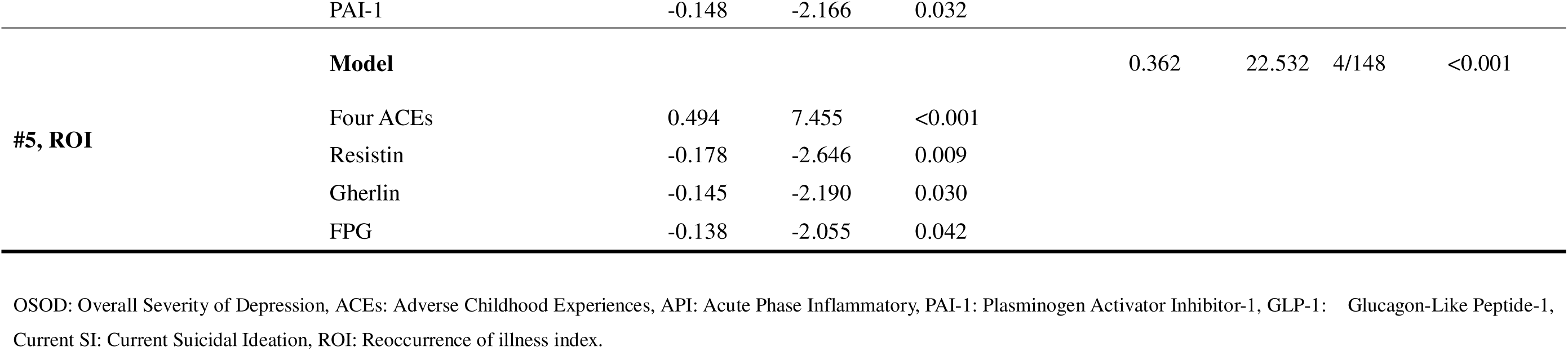
Results of Multiple regression analysis with clinical rating scale scores as dependent variables.

## 4. Discussion

### 4.1. Hormonal and Metabolic Signature in Chinese MDD Patients

This study delineates a distinct neuroendocrine-metabolic profile in Chinese MDD patients, characterized by lower fasting insulin, glucagon, and PAI-1 levels, alongside an elevated API index.

This profile of reduced insulin and PAI-1, in the context of heightened inflammation, appears counterintuitive when viewed against the conventional paradigm linking MDD to IR and elevated pro-inflammatory adipokines, as often reported in Western cohorts^10^. A critical contextual factor is the relatively low prevalence of MetS and obesity in our Chengdu sample, consistent with regional epidemiological data^38^ and our previous report ^6,13^. In populations with high MetS prevalence, metabolic dysregulation (e.g., IR, elevated PAI-1) often co-occurs with and may be amplified by MDD, potentially obscuring the primary hormonal alterations intrinsic to the mood disorder ^5,39^.

### 4.2. Hormonal Dysregulation and the Inflammatory Response

We observed that the suppressed hormonal profile was inversely associated with the elevated API index, a composite measure reflecting a chronic, smoldering inflammatory response^3,6^. This finding underscores chronic mild inflammation as a pivotal mediator of the observed endocrine disturbances.

The inverse association between the API index and PAI-1/adiponectin aligns with established biology whereby systemic inflammation actively suppresses the synthesis and secretion of these metabolic hormones. Pro-inflammatory cytokines, such as IL-6 and TNF-α—which are frequently elevated in MDD—are potent inhibitors of adiponectin gene expression and secretion from adipose tissue ^1,40^. Similarly, while PAI-1 is often viewed as an inflammatory marker itself, its production in visceral adipose tissue and liver can be differentially regulated; sustained, low-grade inflammation as seen in our cohort may disrupt normal regulatory feedback, leading to the observed reduction^18,37^. The significant correlation between lower PAI-1/adiponectin and higher IR index further supports a model where inflammation-driven hormonal suppression contributes to a catabolic or maladaptive metabolic state, even in the absence of classical hyperinsulinemia.

This hormonal suppression extends beyond adipokines. The reduction in fasting insulin and glucagon may also be viewed through the lens of a chronic acute-phase response. The liver, the primary site for synthesizing negative acute-phase reactants (albumin, transferrin) and a key target of inflammatory signaling, may experience a reprioritization of protein synthesis during inflammation^41,42^. Cytokines like TNF-α can directly inhibit hepatic synthesis of albumin and transferrin^43^, and this catabolic milieu may concurrently dampen pancreatic endocrine function or enhance insulin clearance, contributing to lower circulating insulin levels^13^. This state of “inflammatory hypometabolism” is consistent with earlier observations in MDD linking lower visceral protein levels (albumin, transferrin) to anorexia, weight loss, and a malnutrition-inflammation axis^42,44^.

Our findings extend this concept, suggesting that the chronic inflammatory process in MDD orchestrates a coordinated downregulation of anabolic hormones and hemostatic factors. Thus, the observed hormonal profile is not merely a comorbidity but appears integral to the pathophysiology of MDD in this population, directly linked to the API response. This reinforces the concept of MDD as a disorder of NIMETOX pathways, where immune activation exerts downstream disruptive effects on metabolic and endocrine homeostasis ^1,31^.

### 4.3. Hormonal-Metabolic Networks and Clinical Symptomatology

Our third major finding shows significant negative correlations between key hormones (ghrelin, PAI-1, adiponectin), IR indices, and the core clinical constructs examined here. The composite GAP index emerged as the strongest hormonal correlate ^1,31^.

Ghrelin exerts neuroprotective, antidepressant, and anti-inflammatory effects within the brain, modulating the HPA axis and hippocampal neurogenesis^16,17^. Its reduction may therefore attenuate these protective functions. Similarly, lower adiponectin may contribute to increased neuroinflammation and oxidative stress, processes linked to physiosomatic symptoms ^8,19^. The reduction in PAI-1 may reflect a severe catabolic-inflammatory state that dysregulates tissue repair pathways ^18^.

Critically, our multivariate regression models positioned ACEs and the API index as the most consistent and potent positive predictors across all clinical domains. This supports a pathogenic model wherein ACEs might establish a lifelong vulnerability ^28,32^. Within this sensitized framework, subsequent stressors may trigger a chronic smoldering inflammatory response (high API), which in turn suppresses protective hormonal secretion (low GAP). This triple hit—ACEs × Inflammation × Hormonal Deficiency—might drive the phenome of MDD, including overall severity, somatic burden, suicidal ideation, and the tendency towards recurrence ^26,45^. Our findings thus integrate the hormonal dysregulation into the previously established “ACE-ROI-inflammation” axis, illustrating how metabolic hormones serve as both mediators and modulators of this core pathological pathway ^32^.

### 4.4. Towards a Stratified Hormonal-Immune-Metabolic Subtype of MDD

Building upon the distinct profile identified, we propose that the constellation of suppressed anabolic/protective hormones (low GAP index), elevated smoldering inflammation (high API index), and enhanced insulin sensitivity delineates a specific “hormonal-immune-metabolic” (HIM) subtype of MDD, which may be particularly salient in populations with low background prevalence of MetS and obesity, such as our Chengdu cohort. This conceptualization moves beyond a one-size-fits-all model of metabolic dysfunction in depression and instead advocates for a region- and metabolism-stratified understanding of its pathophysiology^6,13^.

The contrast with profiles commonly reported in Western studies is instructive. In populations with high MetS prevalence, MDD is frequently associated with elevated IR, PAI-1, and pentameric CRP—a pattern driven by the superimposition of depressive pathophysiology on a pre-existing state of adiposity-driven meta-inflammation^10,46,47^. In such a context, inflammatory pathways are pre-primed, and adipokine secretion is skewed towards a pro-inflammatory, insulin-resistant state^48,49^. Conversely, in our sample with low MetS prevalence, the inflammatory tone appears to be of a different nature: a chronic, low-grade, “smoldering” response, marked predominantly by a negative acute phase reaction (low albumin, transferrin) and a modest rise in monomeric CRP ^3^. This type of inflammation, potentially linked to persistent low-grade immune activation and catabolism, may create a milieu that paradoxically enhances insulin sensitivity in the short term—a hormetic effect observed in some contexts of mild inflammation^50^—while simultaneously suppressing the synthesis of hormones like adiponectin, PAI-1, and ghrelin.

This stratification has significant implications. It explains the discrepant findings regarding IR and PAI-1 in MDD in the literature and highlights that the “metabolic dysregulation” in MDD is not monolithic. It may manifest as “hyper-metabolic” dysregulation (high IR, high atherogenicity) in high-MetS settings versus a “hypo-metabolic” or catabolic dysregulation (low hormones, high inflammation, preserved insulin sensitivity) in low-MetS settings. Recognizing this HIM subtype is crucial for biomarker interpretation, disease subtyping, and personalized treatment.

### 4.5. Diagnostic and Prognostic Implications

The integrated biomarker model developed in this study—combining the hormonal GAP index, the inflammatory API index, and a history of ACEs achieved an AUC of 0.864, demonstrating that peripheral profiling can effectively distinguish MDD patients from healthy controls. This framework opens avenues for mechanism-informed treatment stratification. For instance, patients with this specific HIM profile might be prioritized for treatments aimed at correcting the catabolic state. Conversely, patients with MDD and comorbid MetS might exhibit a different profile (high pCRP, high PAI-1, high IR) and may benefit more from insulin sensitizers or statins ^51,52^. Thus, the GAP and API indices could serve as objective tools to guide personalized therapeutic decisions.

Future research must validate these biomarkers in longitudinal cohorts to establish their predictive value for treatment response and disease progression.

Intervention studies targeting the inflammation-hormone axis (e.g., with anti-cytokine agents or hormone-modulating drugs) in patients selected based on this profile are a critical next step to translate these mechanistic insights into clinical practice.

## 5. Limitations

This study has several limitations that warrant consideration. First, the cross-sectional design precludes causal inferences regarding the relationships between hormonal dysregulation, inflammation, and clinical symptomatology. The single-center recruitment from southwestern China may limit the generalizability of our findings to other populations with distinct genetic backgrounds, lifestyles, and prevalences of metabolic disorders. Furthermore, although we controlled for major confounders, residual influences from unmeasured variables such as detailed dietary intake, physical activity levels, and precise medication histories cannot be entirely ruled out.

While we statistically controlled for the use of psychotropic medications, and analysis showed no significant effect on the primary biomarkers, potential subclinical or long-term modulation of endocrine axes by these drugs cannot be entirely discounted. Additionally, the hormonal and metabolic assessments were based on single fasting blood samples, which may not capture dynamic secretory patterns or circadian rhythms that are critical for hormones such as ghrelin and cortisol. Future longitudinal studies with larger, multi-center cohorts and more frequent biosampling are needed to validate and extend our findings.

## 6. Conclusions

In summary, this study delineates a distinct hormonal-metabolic signature in Chinese patients with MDD, characterized by suppressed levels of key metabolic hormones (notably PAI-1, adiponectin, and ghrelin) within a context of chronic, smoldering inflammatory response, as indexed by an elevated API score. Critically, this profile is strongly associated with core clinical dimensions of the disorder, including overall severity, physiosomatic burden, and recurrence risk. Our findings extend the NIMETOX model of depression by integrating hormonal and adipokine components, revealing a pathophysiological axis where ACEs, persistent inflammation, and hormonal suppression converge to drive the clinical phenotype.

These results advocate for a stratified, context-dependent understanding of metabolic dysfunction in MDD. The identified HIM subtype may be particularly relevant in populations with low MetS prevalence. The integrative biomarker model (GAP-API-ACEs) demonstrates promising diagnostic and prognostic utility, paving the way for mechanism-informed subtyping and personalized treatment strategies that aim to modulate the inflammation-hormone axis in a targeted subset of patients.

## Supporting information

Electronic Supplementary File

## Data Availability

Data will be available on request.

## Acknowledgements

Not Applicable.

## Ethical approval

This study was approved by the Ethics Committee of Sichuan Provincial People’s Hospital (Approval No. [Ethics (Research) 2024-203]), and written informed consent was obtained from all participants.

