## Supplementary material for "Metabolic Hormone and Adipokine Alterations in Major Depressive Disorder in Relation to the Acute-Phase Inflammatory Response and Early-Life Adversity": Electronic Supplementary File

**Supplementary Table 1. Information on the construction of the clinical domain scores.**

| Current-SI | A principal component extracted from six items of C-SSRS assessing current suicidal ideation intensity and frequency. |
| --- | --- |
| ROI | Z composite score: z (number of depressive episodes) + z (frequency of suicidal ideation) + z (frequency of suicidal attempts). |
| Physiosomatic symptoms | A principal component extracted from three somatic symptom domains which shape the single group factor, namely SSS-8, pure CFS and pure physiosomatic |
| OSOD | Overall severity of depression, constructed as z score of a principal component from vegetative, affective, and physiosomatic symptom domains. |

Current SI: Current Suicidal Ideation; C-SSRS: Columbia Suicide Severity Rating Scale; ROI: Recurrence of Illness; SSS-8: Somatic Symptom Scale-8; CFS: chronic fatigue syndrome; OSOD: Overall Severity of Depression.

**Supplementary Table 2.** Diagnostic performance metrics of different models in distinguishing patients with major depressive disorder from healthy control groups.

| **Predictors** | **AUC** | **SE** | **Gini index** | **Max K-S** | **Overall Model Quality** | **Sensitivity/Specificity** |
| --- | --- | --- | --- | --- | --- | --- |
| Ghrelin, PAI-1, Adiponectin | 0.741 | 0.040 | 0.482 | 0.425 | 0.660 | 0.688/0.615 |
| Ghrelin, PAI-1, FPG | 0.736 | 0.047 | 0.471 | 0.362 | 0.640 | 0.702/0.641 |
| Ghrelin, PAI-1, API index | 0.776 | 0.042 | 0.552 | 0.415 | 0.690 | 0.718/0.667 |
| Ghrelin, PAI-1, API index, sex, Four ACEs | 0.818 | 0.036 | 0.636 | 0.509 | 0.750 | 0.742/0.718 |
| GAP index, API index, Four ACEs | 0.864 | 0.031 | 0.728 | 0.591 | 0.800 | 0.798/0.744 |

PAI-1: Plasminogen Activator Inhibitor-1, API: Acute Phase Inflammatory, ACEs: Adverse Childhood Experiences, GAP: Ghrelin+ Adiponectin+ PAI-1.
